# Event-based seizure detection in human iEEG with neuromorphic hardware

**DOI:** 10.1101/2025.07.10.25331024

**Authors:** Flavia Davidhi, Filippo Costa, Debora Ledergerber, Giacomo Indiveri, Lukas Imbach, Johannes Sarnthein

## Abstract

**Background:** Epilepsy is a neurological disorder that affects approximately 1% of the global population. The current method for seizure monitoring, seizure diaries, is often inaccurate, making precise monitoring challenging. A monotonic descending “chirp” pattern in intracranial EEG (iEEG) is a specific marker of seizure onset and can be used for automatic detection.

**Objective:** To determine whether a spiking neural network (SNN) implemented on the DYNAP-SE1 neuromorphic chip can detect chirps/seizures.

**Methods:** We analysed 48 h of continuous bipolar iEEG from one patient (40 seizures). The signal was filtered into six 10 Hz sub-bands between 0 and 40 Hz, then encoded into UP/DOWN events with software asynchronous delta modulation (ADM). The encoded signal was used as input in the hardware-implemented SNN with 34 adaptive-exponential neurons (3.3 % of the 1024 neurons on one chip). Hierarchical inhibition enforced the required high-to-low band sequence, and a disinhibition unit suppressed isolated low-frequency bursts. We implemented the same 28-neuron SNN (without the dis-inhibition population) on software.

**Results:** Chirps appeared at the onset of all 40 seizures (100 %). The hardware SNN detected every seizure (sensitivity = 100 %) and produced one false alarm in 48 h (falsealarm rate = 0.021 h**^-1^**). Mean processing time was 4 h 55 s ± 42 s for each 4-h data block, showing real-time operation. In the software SNN implementation, we analysed the same 48-h iEEG bipolar channel recording, and detected 32/40 seizures (80 % sensitivity) with 9 false alarms (false-alarm rate = 0.19 h**^-1^**).

**Conclusion:** A 34-neuron SNN implemented on DYNAP-SE1 detects seizures from single-channel iEEG in real time with 100% sensitivity and a low false-alarm rate while using minimal hardware resources.

## 1 Introduction

Epilepsy affects approximately 50 million people worldwide, making it one of the most common chronic neurological disorders [1]. Approximately 30 % of those patients develop drug-resistant epilepsy [2]. Reliable seizure detection is critical for treatment decisions. However, traditional seizure monitoring methods, such as seizure diaries, are often inaccurate. While electrographic seizures are considered the gold standard, expert review of prolonged EEG is burdensome and prone to under-reporting [3]. This highlights the need for precise and automated seizure detection systems. New seizure detection methods using automated EEG analysis are under development to improve accuracy [4].

Monotonic descending frequency patterns (chirps) in the intracranial EEG (iEEG) are strongly associated with seizures [5–10]. In both humans and animal models, a chirp typically manifests as an initial burst of fast oscillations (30–100 Hz) that rapidly decays in frequency [11]. Chirps have been identified as highly specific markers of seizure onset, particularly in focal epilepsies characterized by low-voltage fast activity (LVFA) [11]. Chirps have also been proposed as biomarkers of the epileptogenic zone [8, 12]. Schiff et al. [5] reported observing chirps in 83% of 42 seizures they examined from 6 patients. Additionally, in a monocentric cohort of 105 patients with iEEG recordings, 88.6% exhibited LVFA accompanied by chirp patterns, showing the prevalence of this feature in focal seizures [11].

Automated chirp detection is a potential avenue for seizure detection. Early methods showed feasibility [10]. A recent synthetic chirp dataset has enabled transformer-based chirp detection [13]. Real-time deployment remains computationally intensive, making neuromorphic hardware attractive due to its efficiency and scalability. The DYNAP-SE1 neuromorphic chip [14] offers low-power consumption, event-driven and real-time processing capabilities, making it particularly suited for processing biological signals such as iEEG [15–17]. DYNAP-SE1 implements event-driven computing: it processes data only when spiking activity occurs, resulting in ultra-low power consumption [14]. These features are advantageous for seizure detection systems, which require continuous “always-on” monitoring while maintaining energy efficiency for long-term use (days to months).

Spiking neural networks (SNNs) implemented on neuromorphic hardware have shown robustness in processing iEEG signals, particularly in identifying epileptic biomarkers such as high-frequency oscillations (HFOs). SNNs implemented on the DYNAP-SE1 chip have effectively detected HFOs in iEEG and in intraoperative electrocorticography [15, 17, 18] and seizures [19]. This shows the potential of SNNs for clinical applications that require real-time iEEG processing.

Several SNN-based algorithms can be implemented on DYNAP-SE. For example, the neural state machine (NSM) algorithm was introduced as a novel SNN architecture for classification of spatiotemporal patterns [20]. Subsequent refinements to the NSM network enabled its application to biomedical signal processing, specifically electrocardiogram (ECG) data, detecting monotonic increases in heart rate, while maintaining a low power budget, suitable for wearable devices [21, 22].

In the present study, we aim to explore the detection of seizure patterns using iEEG data, focusing on chirp patterns, using neuromorphic hardware (DYNAP-SE1), building on previous detection of high-frequency oscillation patterns with the same technology [23].

## 2 Results

### 2.1 A chirp pattern in the iEEG initiates the seizure

Figure 1 shows an example of a chirp at seizure onset. The chirp occured in the left hippocampus, which was the seizure onset zone. All chirps were visually observed at seizure onset in 40 out of 40 (100%) seizures. All chirps were observed in the frequency range 0-40Hz, thus we focused our analysis on that frequency range.

**Fig. 1.**
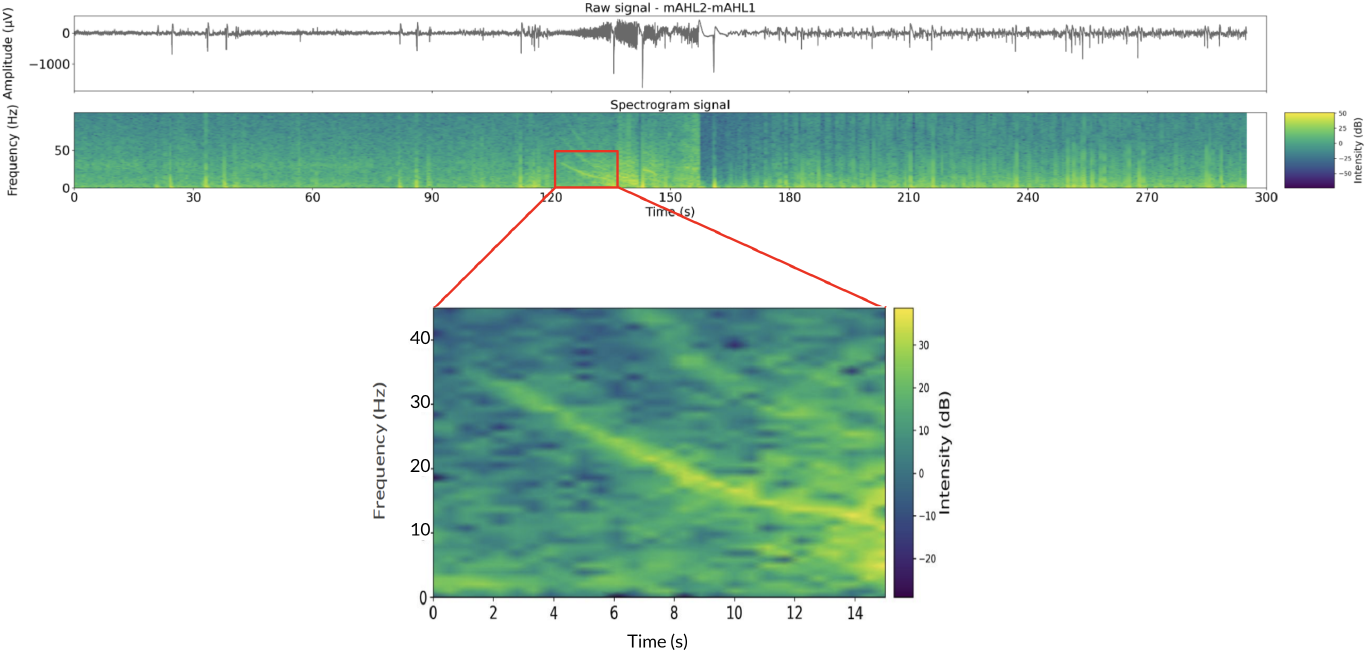
Example of chirp at seizure onset.

### 2.2 Signal preprocessing and encoding

To capture the chirp pattern, we devised and used the pipeline shown in Figure 2. We bandpass filtered the iEEG signal in the bands 10-20Hz, 15-25Hz, 20-30Hz, 25-35Hz and 30-40Hz, and lowpass filtered in the band 0-10Hz. We used 2nd-order IIR (Butterworth) filters, which are compatible with hardware filtering.

**Fig. 2.**
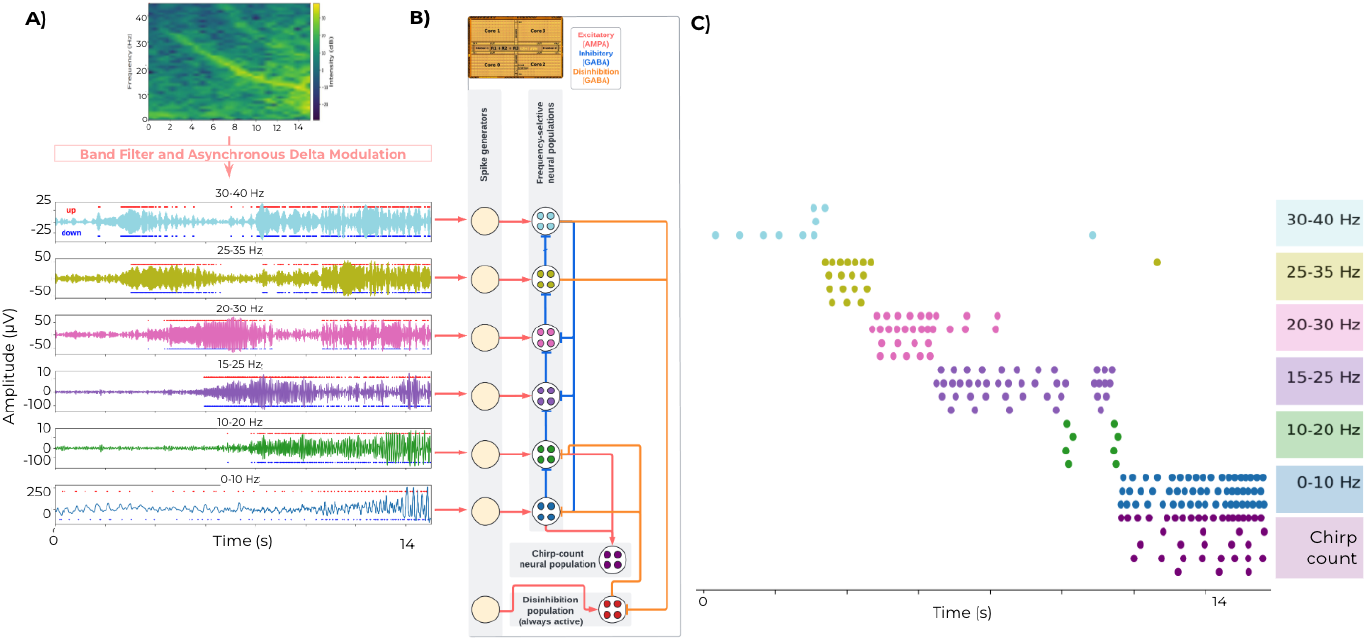
SNN. A) The input signal is iEEG filtered in frequency bands corresponding to the chirp (0-10 Hz, 10-20 Hz, 15-25 Hz, 20-30 Hz, 25-35 Hz, 30-40 Hz). B) The signal filtered in frequency bands is converted into UP and DOWN events using ADM. The UP and DOWN events are sent to the spike generators in the DYNAP-SE1 FPGA (one spike generator per frequency band). C) The SNN features one neural population per frequency band, and one chirp-counting neural population. The SNN employs two kinds of connections: inhibitory (GABA) and excitatory (AMPA). Each frequency band population inhibits all others aside from the previous one (from highest to lowest frequency band).

We employed a software Asynchronous Delta Modulation (ADM) encoder to convert analog waveforms (iEEG) into two streams of UP/DOWN events (Figure 2A). The first 100 seconds of each 4-hour iEEG recording were set as the baseline. Based on this baseline, two thresholds were computed: an UP threshold (baseline mean + 7×baseline standard deviation) and a DOWN threshold (baseline mean -7×baseline standard deviation). Each time the iEEG signal crosses one of these thresholds, an UP or DOWN event is generated, thereby encoding the signal into a stream of UP/DOWN events.

### 2.3 SNN implementation in DYNAP-SE

We devised an SNN that we implemented on the DYNAP-SE neuromorphic chip. The SNN architecture (Figure 2B) consists of three types of neural populations:

- **Frequency band neural populations**: These receive the UP/DOWN events from iEEG filtered frequency bands.
- **Chirp-counting neural population**: This population detects the activation of the two lowest frequency band populations, thus detecting chirps/seizures.
- **Disinhibition neural population**: This modulates the activity of the frequency band populations to reduce false detections.

Each frequency band is assigned a dedicated neural population, resulting in six neural populations per channel. There is a spike generator in the DYNAP-SE FPGA for each frequency band population and for the disinhibition population. These spike generators receive UP/DOWN pulses from the ADM encoding of the input signal. Each spike generator interfaces with its respective neural population.

The neural populations are interconnected hierarchically to ensure chirp detection (Figure 2B). Each frequency-band neural population inhibits all others aside from the next one, in order from high to low frequency. The two lowest frequency populations are connected to the chirp-counting population, which identifies chirps/seizures.

The disinhibition neural population is continuously active. It inhibits the two lowest frequency populations and, in turn, is inhibited by the three highest frequency populations. This connectivity ensures that chirps are detected only when frequency band populations activate in the correct sequence, as shown in Figure 2C, minimising false alarms caused by unrelated low-frequency activity. The SNN is NSM-inspired, as it detects different states (activation of frequency bands), and it counts chirps only if the descending monotonic pattern is detected.

The proposed SNN is compact. It comprises 34 neurons in total. We are currently using 3.3% of DYNAP-SE 1024 silicon neurons in one chip [14], ensuring scalability to more iEEG channels and a future integration into a wearable device. The network is also low-parameter: the SNN is governed by 50 parameters.

### 2.4 The compact hardware SNN detects chirps in real time

With our SNN neuromorphic pipeline, in 48 consecutive hours of one iEEG channel from one patient, we detected all 40 seizures (sensitivity = 100%) with only one false detection (false detection rate = 0.021 per hour), as shown in Figure 3. The iEEG data was processed in 4-hour segments. The AMPA weight parameter in core 2, where the counting neurons are located, was calibrated on one seizure in every four hour recording (mean AMPA synaptic weight fine value: 59.09 ± 9.50). For each 4-hour segment, the mean processing duration was 4 hours and 55 seconds (standard deviation = 41.72 seconds).

**Fig. 3.**
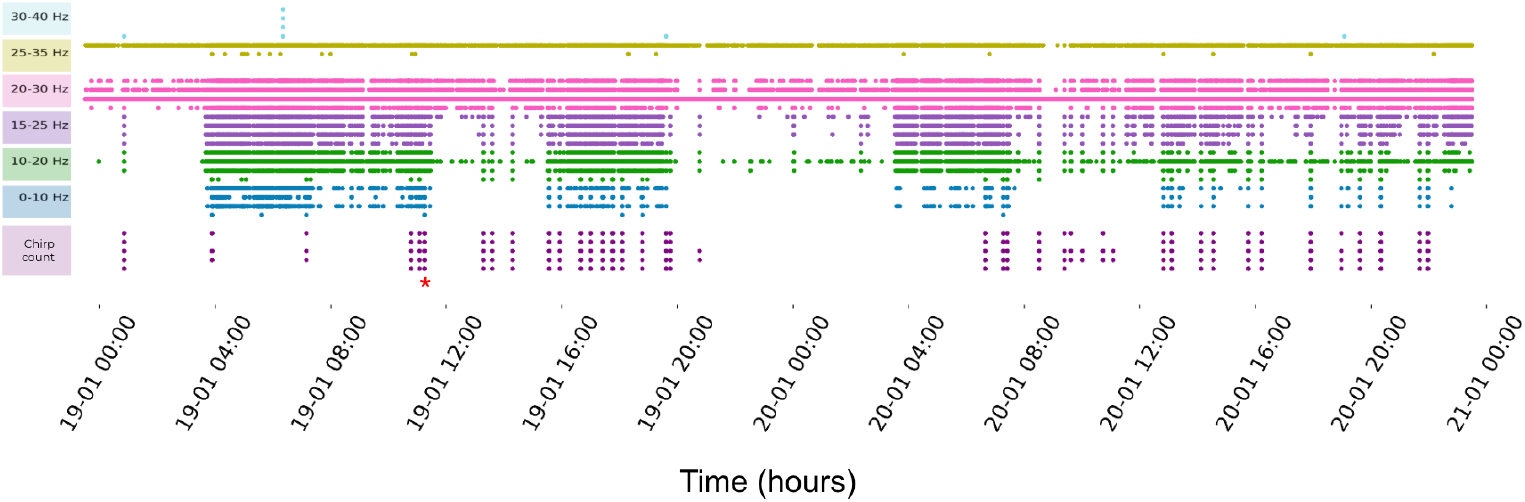
Output activation recorded from the DYNAP-SE neuromorphic chip. In 48 continuous hours of iEEG data, 40 seizures were correctly identified, as shown in the chirp-count section of the graph. One false alarm was recorded, as identified by the red asterisk *.

### 2.5 Software simulation

To verify that the chirp–counting mechanism is sufficient for seizure detection, we ran the same SNN we implemented in hardware (albeit without the disinhibition population) on the same 48-hour iEEG recording (channel = mAHL1-mAHL2). The chirp detection parameters were set to `WIN SEC = 5.0 s, MIN SPIKES = 100, REFRACT SEC = 100 s` (see Methods). An alarm was issued if, in the 20 s preceding it, ≥ 20 events occurred in the two highest-frequency populations and ≥ 20 events occurred in the 5 s preceding it in the second-lowest-frequency population. For each 4-hour segment, the mean processing duration was 16 min 53 s (standard deviation = 9.27 s).

Figure 4 shows the raster plot with the activity recorded from the software SNN. 32 out of 40 seizures were detected, while 9 alarms occurred outside seizure periods. This yields a sensitivity of 80 % and a false-alarm rate of 0.19 h^-1^.

**Fig. 4.**
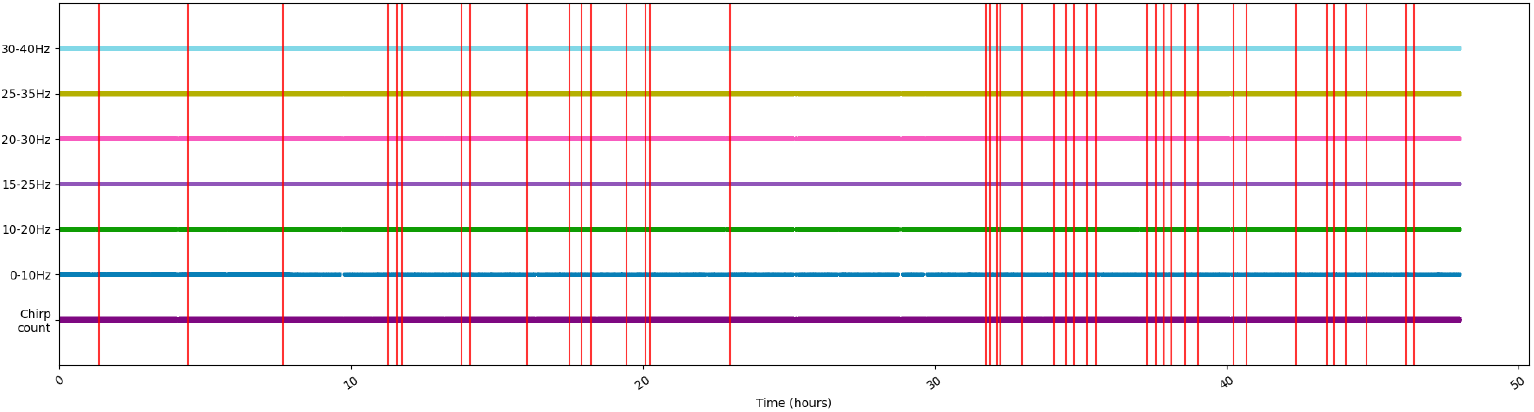
Output activation recorded from the software SNN. In 48 continuous hours of iEEG data, 32/40 seizures were correctly identified, with 9 false alarms. Red vertical bars mark all detections.

## 3 Discussion

We showed that a compact SNN can detect seizures in real-time with high sensitivity (100%) and a low false-alarm rate (0.021 h^-1^ or 0.5 per day) using a single iEEG channel (Figure 3).

### 3.1 Advantages of SNN-based detection

Our SNN does not rely on large-scale supervised training, since it was configured on the patient chirp pattern (using six band-specific filters). By looking at a few examples of chirps in a specific patient, one can determine the pattern and set the filters for each patient accordingly. The ability to perform well with few examples (in this case, we established the ADM thresholds on a short baseline and then detected 40/40 seizures with minimal parameter changes) is an advantage of the presented SNN strategy. It implies that new patients could be onboarded with minimal calibration (just by tuning the filter bank).

Our neuromorphic SNN comprises only 35 neurons (Figure 2) with 50 chip tunable parameters. The entire network uses 3.3% of the 1024 neurons available on a single DYNAP-SE1 chip. This leaves headroom to scale up the system to monitor multiple iEEG channels or to detect additional biomarkers in parallel. Adding more channels would increase resource usage linearly (each additional channel would be handled by another 35 neurons), still within on-chip limits. Indeed, multi-channel SNN deployments have already been demonstrated for similar tasks, such as high-frequency oscillation detection [15–18, 24].

The DYNAP-SE1 chip implements event-driven computation, which means it only uses energy when an input spike occurs [14], without making use of continuous high-rate digital clocked processing. This makes SNN-based detectors feasible to run in an always-on fashion on a battery-powered wearable. Our pipeline processed data in real time with a mean latency of 55s per 4h of iEEG on chip, demonstrating the capability for live seizure monitoring while carrying out computation at the edge.

The architecture is also inherently interpretable. Each neural population in the network has a role (responding to a specific frequency band or counting the chirp occurrence, as shown in Figure 2), and the connectivity enforces known rules (high-to-low frequency order, etc.). This transparency can be advantageous in a clinical setting, where understanding why an alarm was raised (e.g. which frequency bands were activated) builds trust in the system.

### 3.2 Limitations

Our method was evaluated on a single patient’s iEEG; performance in a broader patient cohort remains to be validated. However, given that chirps are a very specific pattern, we expect a low false-positive rate to generalize, provided the patient exhibits chirp-type seizure onsets. An advantage of our SNN is that it encodes a general rule for chirps (descending bands sequence) that could apply out-of-the-box to other patients who manifest chirps, potentially with only minor adjustment of filter bank parameters (e.g. different patients can have chirps at different frequencies). This needs validation in a larger cohort of patients to ensure the algorithm generalises to different brain regions and epilepsy types.

A technical limitation in the current approach is the recalibration of the counting neurons AMPA weight for each 4-hour iEEG recording. We hypothesise that this was needed since we employ a different baseline for the ADM encoding for each 4-hour iEEG recording (the baseline is taken as the first 100-second segment of each 4-hour iEEG recording). Thus, in future improvements of our chirp-detection pipeline, we plan on keeping the baseline fixed to one single iEEG segment. This way, we will calibrate the AMPA weight of the counting neurons only once on one chirp for each patient, to then leave it fixed after the initial calibration. It is also necessary to mention that the rest of the network parameters were kept fixed for all chirp/seizure detections described in this study.

### 3.3 Possible clinical applications of SNN electrographic seizure detection

Reliable real-time seizure detection is of clinical importance in the management of epilepsy. The iEEG analyzed in this study was recorded during pre-surgical evaluation with video-monitoring of the patient. In this intensive monitoring setting, automated seizure detection in real-time might be of help to the staff for annotating seizures.

Another application with iEEG recordings might be the implementation in responsive brain stimulation (RNS). However, iEEG is too invasive for implementation in a large number of patients.

In everyday life, patients are rely on self-reported seizure diaries, which are inherently inaccurate [3]. Inadequate seizure logging can mislead physicians about a patient’s condition, risking suboptimal therapy (e.g. under-treating an active epilepsy or over-medicating a stable patient). For wide-spread use, our SNN chirp detection should be embedded in less invasive recording modalities. Such a future device could monitor from alternative electrode modalities such as subscalp (subcutaneous) EEG implants. Subcutaneous EEG electrodes have recently emerged as a minimally invasive option for ultra-long-term monitoring [25]. These devices can record EEG for months in outpatient settings. Seizure detection on subcutaneous EEG has been reported with ∼97% sensitivity and *>*90% specificity [26]. Subscalp systems are also well tolerated over months and provide continuous data coverage ∼75% of the time on average [26]. The combination of a subcutaneous EEG platform with a low-power neuromorphic detector is thus synergistic. We envision an integrated system where a small subcutaneous electrode (for example, a 2-channel implant placed over the epileptic focus) streams event-encoded EEG to an onboard SNN chip. Such a device could function as a wearable seizure detection device, continuously logging seizures or even issuing wireless alerts to the patient’s smartphone or to caregivers when a seizure is detected.

Regarding the detection of electrographic seizures in real-time, several advantages come to mind. First, the 100% sensitivity achieved by our SNN-based detector in this case study means that no seizures went undetected. In a real-world setting, this level of sensitivity could ensure that even seizures occurring at night or without obvious outward signs are captured. The low false alarm rate we report is important to patient acceptance: if a wearable device frequently gave false alerts, patients might abandon it. As chirp-based detection focuses on a specific seizure onset feature [5, 11, 12], it offers an advantage over detectors that rely on less specific changes (such as heart rate or movement), which can be triggered by benign activities. Second, an automated detection system could alert caregivers or trigger interventions that could be lifesaving by enabling timely assistance, for example, turning a patient on their side, administering rescue medication, or notifying emergency services. Third, seizure detection with our system could be validated in real-time. The patient or caregiver could annotate a false alarm simply by pressing on a smart watch. This would be a way to reduce the false alarm rate in the seizure log. Thus, a chirp-based detector could improve the precision of seizure monitoring. Ultimately, one might test whether similar event-based detectors can be adapted for other patterns of seizure onset or for patients who do not exhibit clear chirps. In patients with multiple seizure types, the SNN could be expanded or paired with additional detectors, for example, a separate module for seizures without chirps. Thanks to the scalability of the neuromorphic SNN within the DYNAP-SE chip, integrating multiple detection circuits would be feasible within the same device.

### 3.4 Conclusion

In conclusion, the present study demonstrates a promising combination of a neurobiological marker (chirp) with neuromorphic computing. We have shown that a small SNN can reliably detect seizures/chirps in one patient. The event-based SNN approach offers a lightweight, efficient, and accurate approach to automated seizure detection. This is a promising direction for wearable and implantable seizure detection devices, where energy and size are important requirements. Clinical translation will require testing in more patients and integrating the technology into a chip suitable for daily use. Both of these are achievable given current trends. For instance, subscalp EEG implants are already in human trials [27], and custom neuromorphic chips for health monitoring are rapidly maturing. This work serves as a proof-of-concept that real-time event-based seizure detection is not only feasible but advantageous. By prioritizing low-power operation, high specificity, and interpretability, we aim to deliver a device that improves the safety and quality of life for people with epilepsy.

## 4 Methods

### 4.1 Patient and recording setup

We recorded iEEG from a patient (F) who was implanted with depth electrodes for presurgical evaluation. Depth electrodes (1.3 mm diameter, 8 contacts of 1.6 mm length, spacing between contact centres 5 mm, ADTech®, www.adtechmedical.com) were implanted stereotactically. The iEEG data was acquired at 4000 Hz sampling frequency with an ATLAS recording system (Neuralynx, www.neuralynx.com). We analysed 48 hours of iEEG from the seizure onset zone in the left anterior hippocampus.

### 4.2 Ethics statement

The ethics committee Kantonale Ethikkommission Zürich gave ethical approval for this work (PB-2017-00094).

### 4.3 Neuromorphic hardware: DYNAP-SE

This work employs the DYNAP-SE neuromorphic chip processor [14], an event-driven mixed-signal analog-digital chip that comprises analog silicon neurons and synapses whose dynamics unfold in biological real-time. The device integrates four processing cores, each with 256 adaptive-exponential integrate-and-fire neurons [28, 29]. Every synapse can be configured as one of four modes: fast excitatory (AMPA), slow excitatory (NMDA), fast inhibitory (GABA A), or slow inhibitory (GABA B). Each neuron carries a 64-entry content-addressable memory that stores the addresses of its presynaptic partners, while peripheral asynchronous digital logic handles spike communication via the address-event representation (AER) protocol [30]. The chip’s asynchronous on-chip and inter-chip routing fabric sustains microsecond-scale latencies and preserves flexible connectivity even under heavy traffic conditions.

### 4.4 Software validation

We implemented the SNN in PyTorch using the `snntorch` toolbox. The network architecture was identical to the hardware implementation except that the disinhibition population and its synapses were ablated. The SNN consists of six 4-neuron LIF populations (one per 10 Hz band) and a 4-neuron chirp-count population, comprising 28 neurons in total. As input events, we used the same data employed as input to the hardware SNN (ADM-encoded into UP/DOWN streams with the same parameters used for the hardware preprocessing). All timing experiments were performed on a MacBook Pro (model `Mac15,10`) equipped with an Apple M3 Max SoC (14-core CPU) and 36 GB of unified memory, running macOS Sequoia 15.3.2.

For the seizure-detection logic, we implemented a fixed-length sliding window over the Chirp-count spike times and raised an alarm when `MIN SPIKES` or more spikes fell inside the last `WIN SEC`. After an alarm the detector was silent for `REFRACT SEC`. For the experiment in Section 2.5 we used:

~~~
WIN_SEC = 5.0 # s
MIN_SPIKES = 100 # spikes
REFRACT_SEC = 100.0 # s
~~~

Filter parameters matched those used for the hardware implementation. ADM thresholds were calculated on 100 s baseline (baseline mean + 7×baseline standard deviation) and kept fixed.

### 4.5 Performance evaluation metrics

- **Sensitivity**. Sensitivity is the fraction of seizures that were detected by the algorithm. A seizure is counted as detected if at least one spike from the SNN chirp count population occurs within 20s of the seizure start. Formally

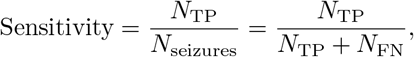

where *N*_TP_ is the number of true–positive seizure detections and *N*_FN_ is the number of non-detected seizures.
- **False alarm rate (FAR)**. The FAR expresses how often the system raises an alarm when no seizure is present. It is given in false positives per hour of recording

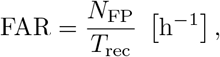

where *N*_FP_ is the number of alarms occurring entirely outside any seizure and *T*_rec_ is the total duration of analysed data in hours.

## Data Availability

All data produced in the present study are available upon reasonable request to the authors.

